# Characterization of bacterial and viral pathogens in the respiratory tract of children with HIV-associated chronic lung disease: a case‒control study

**DOI:** 10.1101/2023.09.11.23295188

**Authors:** Prince K. Mushunje, Felix S. Dube, Jon Ø Odland, Rashida A Ferrand, Mark P. Nicol, Regina E. Abotsi, The BREATHE study team

**Author notes:** Corresponding author: Prince K. Mushunje, MSc, Department of Molecular and Cell Biology, Faculty of Science, University of Cape Town, Cape Town, South Africa. Authors contributed equally (Prince K. Mushunje, Felix S. Dube).

## Abstract

**Introduction:** Chronic lung disease is a major cause of morbidity in African children with HIV infection; however, the microbial determinants of HIV-associated chronic lung disease (HCLD) remain poorly understood. We conducted a case-control study to investigate the prevalence and densities of respiratory microbes among pneumococcal conjugate vaccine (PCV)-naïve children with (HCLD+) and without HCLD (HCLD-) established on antiretroviral treatment (ART).

**Methods:** Nasopharyngeal swabs collected from HCLD+ (defined as forced-expiratory-volume/second<-1.0 without reversibility postbronchodilation) and age-, site-, sex- and duration-of-ART-matched HCLD-enrolled in Zimbabwe and Malawi (BREATHE trial-NCT02426112) were tested for seven bacteria, including *Streptococcus pneumoniae* (SP), *Staphylococcus aureus* (SA), *Haemophilus influenzae* (HI), *Moraxella catarrhalis* (MC), and five viruses, including human rhinovirus (HRV), respiratory syncytial virus A or B, and human metapneumovirus, using qPCR (Fluidigm). Fisher’s exact test and logistic regression analysis were used for between-group comparisons and risk factors associated with common respiratory microbes, respectively.

**Results:** A total of 345 participants (287 HCLD+, 58 HCLD-; median age, 15.5 years [IQR=12.8–18], females, 52%) were included in the final analysis. SP (40%[116/287] *vs.* 21%[12/58], *p* = 0.005) and HRV (7%[21/287] *vs.* 0%[0/58], *p* = 0.032) were more prevalent in HCLD+ patients than in HCLD-patients. Viruses (predominantly HRV) were detected only in HCLD+ participants. HI (1.55×10^4^ CFU/ml *vs.* 2.55×10^2^ CFU/ml, *p* = 0.006) and MC (1.14×10^4^ CFU/ml *vs.* 1.45×10^3^ *CFU/*ml*, p* = 0.031) densities were higher in HCLD+. Bacterial codetection (≥ any 2 bacteria) was higher in the HCLD+ group (36% [114/287] *vs.* (19% [11/58]), (*p* = 0.014), with SP and HI codetection (HCLD+: 30% [86/287] *vs.* HCLD-: 12% [7/58], *p* = 0.005) being the most frequent. In 128 SP-positive participants (116 HCLD+, 12 HCLD-), 66% [85/128] of participants had non-PCV-13 serotypes detected. Serotypes 13 and 21 (9% [8/85] each) and PCV-13 serotypes (4, 19A, 19F: 16% [7/43] each) were more prevalent. Study participants with a history of previous tuberculosis treatment were more likely to carry SP or HI, while those who used ART for ≥2 years were less likely to carry HI and MC.

**Conclusion:** Children with HCLD+ were more likely to be colonized by SP and HRV and had higher HI and MC bacterial loads in their nasopharynx. The role of SP, HI, and HRV in the pathogenesis of CLD, including how they influence the risk of acute exacerbations, should be studied further.

## INTRODUCTION

In 2019, over 2.8 million children and adolescents were living with HIV globally, 90% in sub-Saharan Africa[1]. Respiratory infections remain the most common manifestation of HIV among these children and adolescents [2, 3]. The scale-up of antiretroviral therapy (ART) has increased survival so that growing numbers of children are entering adulthood. In addition, ART has resulted in a reduction in the rate of respiratory disorders, including tuberculosis and lymphocytic interstitial pneumonitis [4–7]. However, studies in sub-Saharan Africa revealed that approximately 30% of HIV-infected older children experience chronic respiratory symptoms, including chronic cough and reduced tolerance to exercise, which often leads to presumptive tuberculosis treatment [8]. The clinical and radiological picture of this chronic lung disease is consistent with small airway disease, predominantly constrictive obliterative bronchiolitis [9].

The pathogenesis of this condition is incompletely understood. It is speculated that HIV-induced chronic inflammation and dysregulated immune activation may play a role [10–12]. A previous study of older children with HIV-associated chronic lung disease (HCLD) conducted by our group demonstrated that there was increased inflammatory activation in children with HCLD (HCLD+) compared to their HIV-infected counterparts without HCLD (HCLD-) [13]. In the same cohort, there was an association between the carriage of specific bacteria in the nasopharynx and HCLD [14]. Specifically, we observed that older children with HCLD were more likely to be colonized with *Streptococcus pneumoniae* (SP) and *Moraxella catarrhalis* (MC) than their HCLD-counterparts [14]. The study utilized bacterial culture, which is limited by viability and a narrow spectrum of culturable bacterial species. Although we observed that SP was associated with HCLD, we did not investigate the specific serotypes that may be involved in this condition, which is important to inform pneumococcal immunization. Carriage of respiratory viruses was also not studied.

Viruses facilitate bacterial infections in the host through various mechanisms, including damaging the respiratory epithelium, modifying the immune response, and altering cell membranes [15]. Coinfection of viruses and bacteria leads to increased bacterial load, thus making individuals more susceptible to complications related to upper respiratory tract infections [16]. Prior to COVID-19, respiratory syncytial virus, influenza virus and human rhinovirus (HRV) were the most common causative agents of upper respiratory infection and have been linked to exacerbations of COPD [17, 18], asthma development [17] and severe bronchiolitis in children [19–21].

To overcome these limitations, we investigated the prevalence of respiratory pathogens in both HCLD+ and HCLD-participants using real-time quantitative polymerase chain reaction (qPCR) to detect and quantify a large number of bacterial and viral targets and elucidate common SP serotypes. We also assessed clinical and sociodemographic factors associated with microbial carriage and density.

## MATERIALS AND METHODS

### Study design, population, and setting

This case‒control study was nested within the BREATHE trial (ClinicalTrials.gov Identifier: NCT02426112) investigating whether azithromycin therapy could improve lung function and reduce the risk of exacerbations among children with HCLD [22]. BREATHE was a two-site, double-blinded, placebo-controlled, individually randomized trial conducted in Harare (Zimbabwe) and Blantyre (Malawi). The study setting, population, and trial procedures are described elsewhere [22–24]. Briefly, we enrolled perinatally HIV-infected participants aged 6 - 19 years with HCLD. HCLD was defined as a forced expiratory volume in 1 second (FEV1) *z* score < −1, with no reversibility (<12% improvement in FEV1 after salbutamol 200 µg inhaled using a spacer) [22]. A group of perinatally HIV-infected children without HCLD (FEV1 *z* score > 0) was also recruited at the same time as the enrollment of trial participants using frequency matching for site, sex, and age and duration of ART to serve as a comparison group for pathogenesis studies. Both groups were on ART for at least six months. All participants were most likely not vaccinated due to the introduction of PCV13 in 2012 in Zimbabwe [25] and in Malawi in 2011 [26], making them ineligible for vaccination at that time because of their older age. Sociodemographic data and clinical history were recorded through an interviewer-administered questionnaire.

### Nasopharyngeal swab collection

Nasopharyngeal swabs were collected at baseline from all participants using sterile flocked flexible nylon swabs (Copan Italia, Brescia, Italy). Swabs were immediately immersed in 1 mL PrimeStore® Molecular Transport Medium (MTM) (Longhorn Vaccines & Diagnostics LLC, Bethesda, USA), transported on ice and stored at −80°C at the diagnostic laboratory at each site. PrimeStore® MTM was used because it is a medium optimized for transporting and storing samples for molecular analyses; it also inactivates potential pathogens and stabilizes nucleic acids [27]. The samples were batched and transported on dry ice to Cape Town, South Africa, where they were stored at −80°C until further processing.

### Total nucleic acid extraction

Total nucleic acid extraction for microbial identification was conducted on NP swabs stored in Primestore® MTM. Briefly, the samples were thawed and vortexed for 10 s, and 400 µl aliquots were transferred into ZR BashingBead^TM^ Lysis Tubes containing 0.5 mm beads (catalog no. ZR S6002–50, Zymo Research Corp., Irvine, CA, United States) for the mechanical lysis steps. Lysis was conducted on a Qiagen Tissue lyser LTTM (Qiagen, FRITSCH GmbH, Idar-Oberstein, Germany) for 5 minutes at 50 Hz, followed by centrifugation (Eppendorf F-45-30-11, Merck KGaA, Darmstadt, Germany) for 1 minute at 10,000 rpm (10,640 g). The supernatants (250 µl) were extracted using the QIAsymphony® DSP Virus/Pathogen Kit (Qiagen GmbH, Hilden, Germany) on the QIAsymphony SP/AS instrument (Qiagen GmbH, Hilden, Germany) following the manufacturer’s instructions. The TNA was eluted in 70 µl DNA elution buffer into the Elution Microtube (Qiagen GmbH, Hilden, Germany) and immediately stored at −80°C until further analysis.

### Real-time qPCR using the Biomark HD system (Fluidigm assay)

The Fluidigm assay was performed at the WITS-VIDA Research Unit, Witwatersrand University, Johannesburg, South Africa. The four bacteria we previously cultured (SP, HI, MC and *Staphylococcus aureus* [SA]), SP and HI serotypes, and viruses (respiratory syncytial virus, human rhinovirus, influenza A and B, human parainfluenza 1 and 3, and human metapneumovirus) were quantified from the total nucleic acid extract using the fluidigm assay previously described by Olwagen *et al.* [28]. These microbial targets were selected on the basis of their possible involvement in HCLD and because they are the most common pathobionts in the nasopharynx. A detailed list of these microbial targets can be found in the supplementary material (Table S1). Data analysis was performed using manually defined thresholds using the Real-Time PCR Analysis Software in the BIOMARK instrument (Fluidigm Corporation, CA, USA). Negative samples were defined as those with Cq values ≥ 37 for each target.

### Data management and statistical analysis

Clinical and sociodemographic data were electronically captured using Google NexusTM tablets (Google, Mountain View, CA, USA) running OpenDataKit software, managed on Microsoft Access databases (Microsoft, Redmond, WA, USA) and analyzed using Stata ((StataCorp, College Station, TX). Comparisons between groups were performed with t tests or Mann–Whitney U tests for continuous data and chi-squared or Fisher’s exact tests for categorical data where appropriate. Multivariate logistic regression, adjusting for age category, duration of ART, site, sex, height-for-age, HIV viral suppression, history of TB treatment, Medical Research Council dyspnea score and ART regimen, was used to investigate the factors associated with microbial carriage and density. The following were excluded from the multivariate model because of colinearity: Enrollment BMI-for-age z score, weight-for-age z score, and CD4 count. A *p* value of less than 0.05 was considered statistically significant.

## RESULTS

### Clinical and sociodemographic characteristics

The study included 345 participants, HCLD+ (n=287) and HCLD-(n=58), with a median age [IQR] of 15.5 (12.8 – 18.0) years and 52% (180/345)] female (Table 1). The median BMI-for-age-*z* score for the HCLD+ group was lower than that for the HCLD- group (−1.1 *vs* −0.4), *p* < 0.001. A higher proportion of the participants from the HCLD+ group were previously treated for tuberculosis (31% *vs.* 12%, *p* =0.001), stunted (49% *vs*. 29%, *p* = 0.009) and underweight (52% *vs*. 14%, *p* <0.001) compared to the HCLD- group. More HCLD+ participants were on a second-line ART (protease inhibitor-based) regimen (25% *vs.* 10%, *p* = 0.01). Ten percent of HCLD+ participants compared to HLCD- (2%) participants had an MRC dyspnea score of 3 or above. None of the participants reported smoking.

**Table 1.** Baseline characteristics of participants in the HCLD+ and HCLD- groups.

| Characteristics |  | HCLD+ [% (n/N)] | HCLD- [% (n/N)] | P<sup>b</sup> |
| --- | --- | --- | --- | --- |
| **Sociodemographic** |  |  |  |  |
| Site | Zimbabwe, % (n/N) | 72% (208/287) | 76% (44/58) | 0.746 |
|  | Malawi | 28% (79/287) | 24% (14/58) |  |
| Sex | Male | 51% (147/287) | 31% (18/58) | **0.006** |
|  | Female | 49% (140/287) | 69% (40/58) |  |
| Age (years) | Median (IQR) | 15.4 (12.9 - 19.9) | 15.7 (12.4 - 18.1) | 0.861 |
| Age groups | 6 - 12 years | 26% (74/287) | 29% (17/58) | 0.373 |
|  | 13 - 16 years | 43% (123/287) | 31% (18/58) |  |
|  | 17 - 19 years | 31% (90/287) | 40% (23/58) |  |
| Currently attending school |  | 81% (231/284) | 86% (50/58) | 0.454 |
| Same age as most children in class <sup>a</sup> |  | 34% (79/233) | 26% (13/50) | 0.442 |
| Repeated $\geq 1$ school grade, | | 58% (162/281) | 40% (23/58) | **0.014** |
| **Anthropometric:** |  |  |  |  |
| BMI for age-z—score | Median (IQR) | -1.1 (-1.8, -0.2) | -0.14 (-0.8, 0.7) | **<0.001** |
| Height for age-z score | <- 2 (Stunted) | 49% (140/287) | 29% (17/58) | **0.009** |
| Weight for age-z score | <-2 (Underweight) | 52% (148/287) | 14% (8/58) | **<0.001** |
| **Current Drugs:** |  |  |  |  |
| Taking cotrimoxazole prophylaxis <sup>a</sup> |  | 91% (259/285) | 87% (48/55) | 0.454 |
| Antiretroviral Regimen | NNRT-base-1st line | 75% (214/287) | 90% (52/58) | **0.010** |
|  | PI-base 2nd line | 25% (73/287) | 10% (6/58) |  |

| **HIV Clinical parameters** |  |  |  |  |
| --- | --- | --- | --- | --- |
| Age at HIV diagnosis (years), | median (IQR) | 7.9 (4.4 - 10.5) | 7.1 (4.1 - 9.8) | 0.377 |
| Age at ART initiation (years), | median (IQR) | 8.5 (5.9 - 11.7) | 8.4 (5.0 - 11.1) | 0.398 |
| Duration on ART (years), | median (IQR) | 6.4 (3.9 - 8.6) | 7.3 (4.7 - 9.0) | 0.219 |
| Duration on ART<sup>a</sup> | > 6 months to <2 years | 9% (26/280) | 3% (2/58) | 0.341 |
|  | 2 to <4 years | 17% (48/280) | 21% (12/58) |  |
|  | 4 to < 6years | 21% (58/280) | 16% (9/58) |  |
|  | 6years or more | 53% (148/280) | 60% (35/58) |  |
| CD4 count categories (Cells/mm<sup>3</sup>) | < 200 | 12% (33/287) | 10% (6/58) | 0.687 |
|  | 200-500 | 29% (84/287) | 24% (14/58) |  |
|  | >500 | 59% (170/287) | 66% (38/58) |  |
| Viral load (VL) suppression<sup>a</sup> | VL < 1000 copies/mL | 54% (153/285) | 66% (38/58) | 0.191 |
| **Respiratory status** |  |  |  |  |
| Hospitalization for chest problems in the last 48 weeks |  | 2% (6/287) | 0% (0/58) | 0.595 |
| Previously treated for TB<sup>a</sup> |  | 31% (88/286) | 12% (7/58) | **0.001** |
| Has asthma<sup>a</sup> |  | 3% (9/286) | 0% (0/58) | 0.366 |
| FEV1 z score | Median (IQR) | -1.9 (-2.46, -1.47) | 0.61 (0.26 - 0.83) | **<0.001** |
| MRC dyspnea score<sup>a</sup> | 1 | 54% (156/287) | 81% (46/57) | **0.006** |
|  | 2 | 36% (103/287) | 17% (10/57) |  |
|  | 3 | 6% (18/287) | 2% (1/58) |  |
|  | 4 | 3% (8/287) | 0% (0/57) |  |
|  | 5 | 1% (2/287) | 0% (0/57) |  |

Abbreviations: HCLD, HIV-associated chronic lung disease; TB, tuberculosis, FEV1, forced expiratory volume in 1 second; IQR, interquartile range; MRC, Medical Research Council; <sup>a</sup> Participants with missing responses were excluded from that variable; *p*<sup>b</sup>, Fisher's exact test

### Prevalence and densities of selected nasopharyngeal microbes in participants with and without HCLD

The prevalence and median densities of selected microbes detected in the nasopharynx of HCLD+ and HCLD-participants are summarized in Table 2. In the HCLD+ group, there was a higher prevalence of SP (40% [116/287] *vs*. 21% [12/58], *p* = 0.005), along with significantly greater median bacterial loads for HI (1.55 × 10^4^ CFU/ml *vs*. 2.55 ×10^2^ CFU/ml, *p* = 0.006) and MC (1.14 ×10^4^ CFU/ml *vs*. 1.45 ×10^3^ CFU/ml, *p* = 0.031). No significant differences were observed in the prevalence and densities of other bacterial species tested. There was a low prevalence of the viruses tested, with HRV (7% [21/287] *vs*. 0% [0/58], *p* = 0.032) detected in the HCLD+ group only. The bacterial species *Klebsiella pneumoniae*, *Neisseria meningitidis*, *Actinobacter baumanii*, *Bordetella pertussis/holmesii*, and viruses influenza A, influenza B, human parainfluenza type 1 & 3, and human metapneumovirus were not detected in any participants.

**Table 2.** The prevalence and density of microbes present in the nasopharyngeal swabs of HIV-infected children with and without HCLD.

| Microorganism | Microbial prevalence |  |  | Microbial density (CFU/ml) |  |  |
| --- | --- | --- | --- | --- | --- | --- |
| | HCLD+ group (n =287) | HCLD- group (n =58) | $p^a$ | HCLD+ group (Median) | HCLD- group (Median) | $p^b$ |
| **Bacterial species** |  |  |  |  |  |  |
| *Streptococcus pneumoniae* (SP) | 40% (116) | 21% (12) | **0.005** | 6.41 x 10<sup>4</sup> | 1.26 x 10<sup>4</sup> | 0.259 |
| *Staphylococcus aureus* (SA) | 6% (17) | 5% (3) | 1.000 | 3.07 x 10<sup>2</sup> | 1.50 x 10<sup>2</sup> | 0.773 |
| *Moraxella catarrhalis* (MC) | 26% (75) | 16% (9) | 0.095 | 1.14 x 10<sup>4</sup> | 1.45 x 10<sup>3</sup> | **0.031** |
| *Haemophilus influenzae* (HI) | 43% (124) | 33% (19) | 0.147 | 1.55 x 10<sup>4</sup> | 2.55 x 10<sup>2</sup> | **0.006** |
| *Haemophilus influenzae* type B | 0% (0) | 2% (1) | 1.000 | - | *1.01 x 10<sup>3</sup> | - |
| *Streptococcus oralis* | 3% (8) | 2% (1) | 1.000 | 8.34 x 10<sup>2</sup> | *2.14 x 10<sup>3</sup> | 0.699 |
| *Neisseria lactamica* | 2% (6) | 0% (0) | 0.595 | 1.51 x 10<sup>3</sup> | - | - |
| *Streptococcus pyogenes* | 2% (5) | 0% (0) | 0.594 | 2.54 x 10<sup>6</sup> | - | - |
| **Virus** |  |  |  |  |  |  |
| Respiratory syncytial virus A | 0.4% (1) | 0% (0) | 1.000 | * 2.74 x 10<sup>2</sup> | - | - |
| Respiratory syncytial virus B | 0.4% (1) | 0% (0) | 1.000 | * 62 | - | - |
| Human rhinovirus | 7% (21) | 0% (0) | **0.032** | 1.35 x 10<sup>4</sup> | - | - |

CLD =Chronic lung disease; <sup>a</sup>Fisher's exact test; <sup>b</sup> Mann–Whitney U test, \*Actual microbial density obtained from one participant

### Nasopharyngeal bacteria and viruses cocolonization in participants with and without HCLD

Bacteria and virus cocolonization detected in HIV-infected participants with or without HCLD is summarized in Table 3 and Table S2. Bacterial detection (any) was significantly higher in HCLD+ (61% [175/287]) than in HCLD-(43.1% [25/58]) (*p* = 0.013). Moreover, the codetection of multiple bacterial species was higher in the HCLD+ group (35.9% [103/287]) than in the HCLD- group (19% [11/58]) (*p* = 0.014). The most frequent bacteria codetected with SP were HI (HCLD+: 33.8% [97/287]) *vs.* HCLD-: 17.2% [10/58], *p* = 0.013) and MC (HCLD+: 23.3% [67/287] *vs.* HCLD-: 12.1% [7/58], *p* = 0.078). Viruses were detected only in the HCLD+ group (8% [23/287]), with virus and bacteria cocolonization reported in 6.6% (19/287) of HCLD+ participants.

**Table 3.** Bacteria and virus cocolonization in study participants at baseline.

|  | HCLD+ group<br>% (n/N) | HCLD - group<br>% (n/N) | *P*<sup>a</sup> |
| --- | --- | --- | --- |
| **Bacterial detection** |  |  |  |
| Single bacterial species | 61% (175/287) | 43.1% (25/58) | **0.013** |
| At least 2 bacterial species | 35.9% (103/287) | 19% (11/58) | **0.014** |
| > 2 bacterial species | 21.3% (61/287) | 12.1% (7/58) | 0.147 |
| **Bacterial cocolonization** |  |  |  |
| SP & SA | 3.1% (9/287) | 5.2% (3/58) | 0.432 |
| SP & HI | 33.8% (97/287) | 17.2% (10/58) | **0.013** |
| SP & MC | 23.3% (67/287) | 12.1% (7/58) | 0.078 |
| SA & HI | 2.1% (6/287) | 1.7% (1/58) | 1.000 |
| SA & MC | 1.1% (3/287) | 3.5% (2/58) | 0.198 |
| MC & HI | 18.8% (54/287) | 12.1% (7/58) | 0.261 |
| SP & other bacterial species <sup>a</sup> | 4.9% (14/287) | 3.5% (2/58) | 1.000 |
| MC & other bacterial species <sup>a</sup> | 3.1% (9/287) | 1.7% (1/58) | 1.000 |
| HI & other bacterial species <sup>a</sup> | 5.2% (15/287) | 3.5% (2/58) | 0.748 |
| SA & other bacterial species | 3.1% (9/287) | 0% (0/58) | 0.366 |
| **Virus detection** |  |  |  |
| Single virus | 8.0% (23/287) | 0% (0/58) | **0.019** |
| ≥ 2 viruses | - | - | - |
| **Viruses and bacteria codetection** |  |  |  |
| Single virus + single bacterial species | 6.6% (19/287) | 0% (0/58) | 0.053 |
| Single virus + multiple bacterial species | 5.6% (16/287) | 0% (0/58) | 0.084 |

Other bacteria <sup>a</sup>; bacteria analyzed as one group (*Klebsiella pneumoniae*, *Neisseria meningitidis*, *Actinobacter baumannii* and *Bordetella pertussis/holmesii*, HI type B, *S. oralis*, *N. lactamica* & *S. pyogenes*); *p*<sup>a</sup>, Fisher's exact test

### SP serotypes and their density detected at baseline in children with and without HCLD

Pneumococcal serotype was assigned for 150 pneumococci from 128 SP-positive samples, including 134 from the HCLD+ group and 16 from the HCLD- group (Figure 1). Carriage with multiple serotypes was detected in HCLD+ (14% [16/116] and HCLD-(17% [2/12]) of the SP-positive participants. The serotype distribution between the HCLD+ and HCLD- groups showed no statistically significant difference, with non-PCV13 serotypes being more prevalent [HCLD+: 69% (93/134) *vs* HCLD-: 81% (13/16)]. The prevalence of PCV-13 serotypes in the HCLD+ group was 31% (41/134) compared to the HCLD- group [19% (3/16)] (*p* =0.398). Among the PCV-13 serotypes, serotypes 4 (16% [7/44]), 19F (16% [7/44]), 19A (16% [7/44]) and 18C (14% [6/44]) were the most prevalent in both groups, while serotypes 13 and 21 accounted for 8% (8/106) each among the non-PCV-13 serotypes. There was no statistically significant difference in median densities between the PCV-13 and non-PCV 13 serotypes (Figure S1). The overall median serotype density was 8.8 CFU/ml.

**Figure 1.**
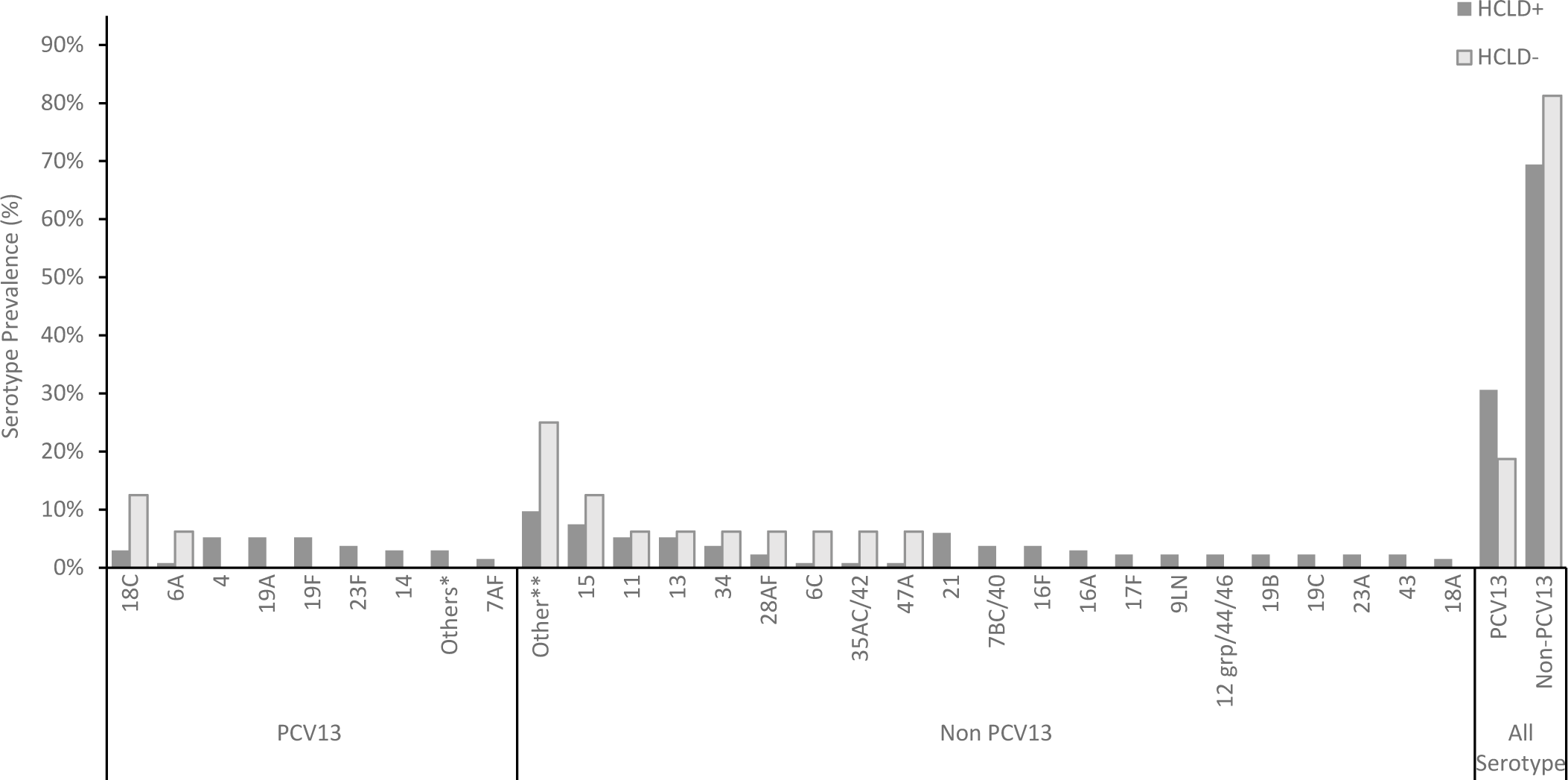
Pneumococcal serotypes recovered from nasopharyngeal swabs of HCLD+ and HCLD-patients at baseline. Abbreviations: PCV, polysaccharide-conjugated vaccine; n, number of isolates serotyped using the fluidigm assay at baseline from the HCLD+ group (n = 116) and HCLD- group (n = 12) Others* HCLD+ group: PCV13 serotype [3 (0.7%), 5 (0.7%), 6B (0.7%), 9AV (0.7%), 4 (5.2%), Others**: HCLD+ group non-PCV 13 serotype [18B (0.7%), 19 atypical (0.7%), 20 (0.7%), 23B (1.5%), 27 (0.7%), 25AF/38 (0.7%), 45 (0.7%), 29 (0.7%), 31 (0.7%), 33C (1.5%), 38 (0.7%)]; HCLD- group non-PCV13 serotype [10A (6.3%), 10B (6.3%), 22A (6.3%), 33B (6.3%)] 15: HCLD+ group non-PCV 13 serotype [15AF (3.7%), 15BC (2.2%), 15like (1.5%)]; HCLD- group non-PCV13 serotype [15like (6.3%)] 11: HCLD+ group non-PCV 13 serotype [11AD [3.7%), 11E (1.5%)]; HCLD- group non-PCV13 serotype [11E (6.3%)]

### Factors associated with carriage of selected bacteria at baseline in participants with HCLD

The results of the univariate and multivariate analyses of the clinical and sociodemographic factors associated with the carriage of SP and SA are displayed in Table 4, while those for HI and MC are shown in Table 5. On multivariate analysis, participants previously treated for TB (adjusted odds ratio were more likely to carry SP (aOR): 2.1 [1.2 −3.9], *p* = 0.016) or HI (aOR: 2.1 [1.2 – 3.8], *p* = 0.011). Participants on ART for ≥2 years (aOR: 0.3 [0.1 – 0.8], *p* = 0.016) and living in Zimbabwe (aOR: 0.5 [0.3 – 0.9], p = 0.014) were less likely to carry HI (Table 5). Similarly, MC carriage was less likely in participants who had been on ART for ≥ 2 years (aOR: 0.4 [0.1 – 0.9], *p* = 0.024) (Table 5). Participants who were attending school were more likely to carry MC (aOR: 2.5 [1.0 −6.2], *p* = 0.047) (Table 5). Female participants were less likely to carry SA at baseline (aOR: 0.3 [0.1 – 1.0], *p* = 0.046) (Table 4).

**Table 4.**
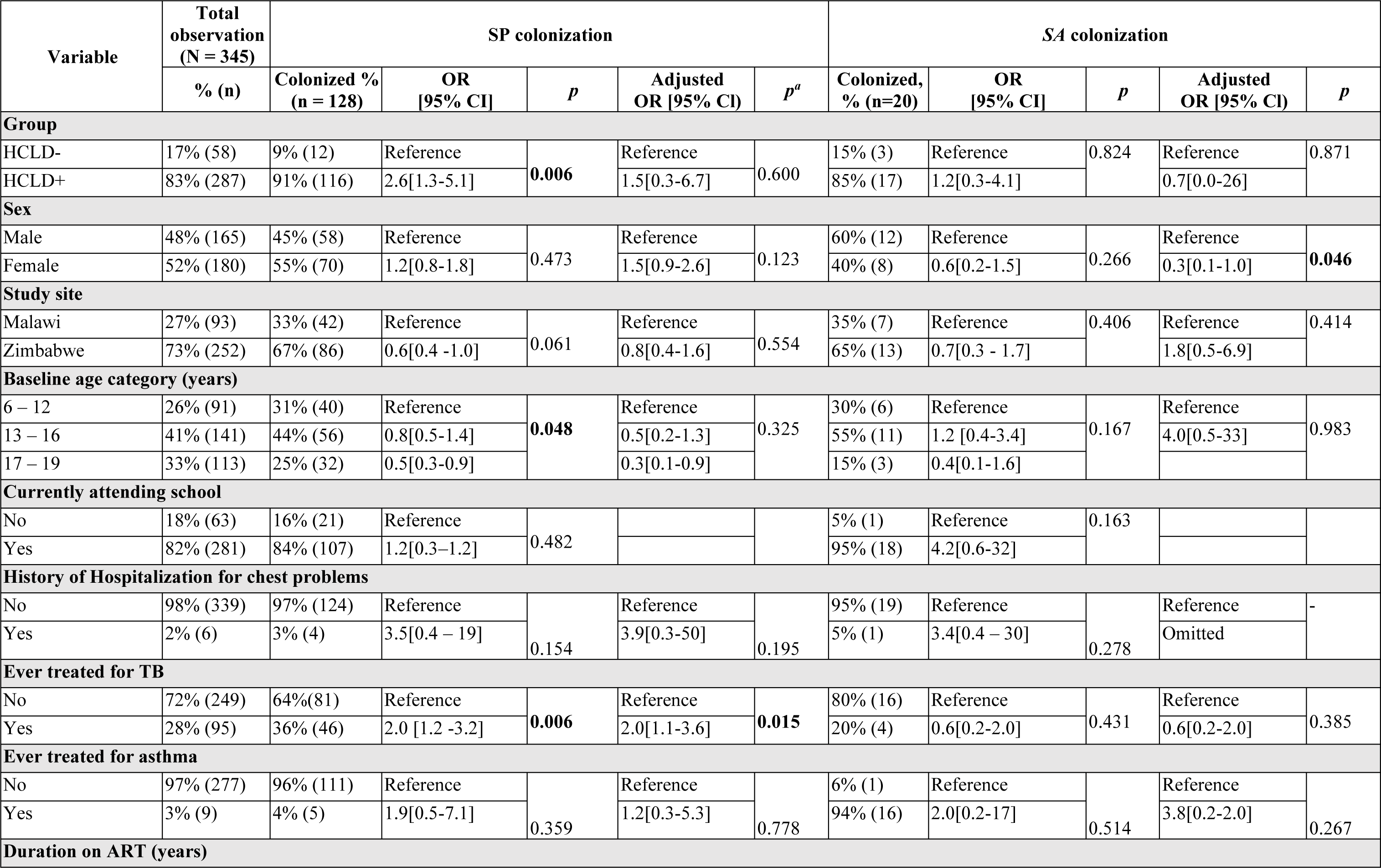
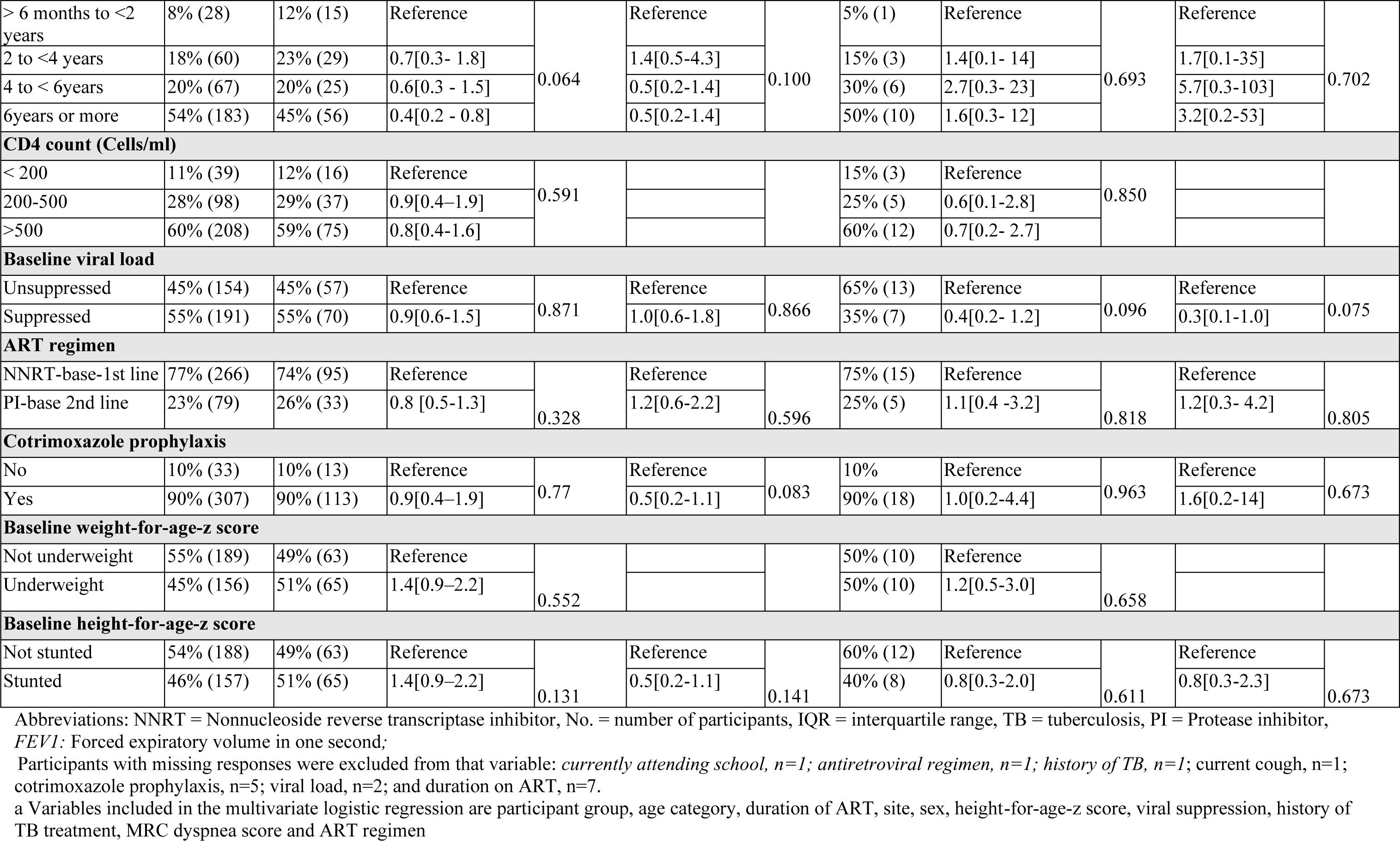
Univariate and multivariate analysis of factors associated with nasopharyngeal SP and SA colonization at baseline.

| Variable | Total observation<br>(N = 345) | SP colonization |  |  |  |  | SA colonization |  |  |  |  |
| --- | --- | --- | --- | --- | --- | --- | --- | --- | --- | --- | --- |
|  | % (n) | Colonized %<br>(n = 128) | OR [95% CI] | p | Adjusted OR [95% CI] | p<sup>a</sup> | Colonized,<br>% (n=20) | OR [95% CI] | p | Adjusted OR [95% CI] | p |
| **Group** |  |  |  |  |  |  |  |  |  |  |  |
| HCLD- | 17% (58) | 9% (12) | Reference | **0.006** | Reference | 0.600 | 15% (3) | Reference | 0.824 | Reference | 0.871 |
| HCLD+ | 83% (287) | 91% (116) | 2.6[1.3-5.1] |  | 1.5[0.3-6.7] |  | 85% (17) | 1.2[0.3-4.1] |  | 0.7[0.0-26] |  |
| **Sex** |  |  |  |  |  |  |  |  |  |  |  |
| Male | 48% (165) | 45% (58) | Reference | 0.473 | Reference | 0.123 | 60% (12) | Reference | 0.266 | Reference | **0.046** |
| Female | 52% (180) | 55% (70) | 1.2[0.8-1.8] |  | 1.5[0.9-2.6] |  | 40% (8) | 0.6[0.2-1.5] |  | 0.3[0.1-1.0] |  |
| **Study site** |  |  |  |  |  |  |  |  |  |  |  |
| Malawi | 27% (93) | 33% (42) | Reference | 0.061 | Reference | 0.554 | 35% (7) | Reference | 0.406 | Reference | 0.414 |
| Zimbabwe | 73% (252) | 67% (86) | 0.6[0.4 -1.0] |  | 0.8[0.4-1.6] |  | 65% (13) | 0.7[0.3 - 1.7] |  | 1.8[0.5-6.9] |  |
| **Baseline age category (years)** |  |  |  |  |  |  |  |  |  |  |  |
| 6 – 12 | 26% (91) | 31% (40) | Reference | **0.048** | Reference | 0.325 | 30% (6) | Reference | 0.167 | Reference | 0.983 |
| 13 – 16 | 41% (141) | 44% (56) | 0.8[0.5-1.4] |  | 0.5[0.2-1.3] |  | 55% (11) | 1.2 [0.4-3.4] |  | 4.0[0.5-33] |  |
| 17 – 19 | 33% (113) | 25% (32) | 0.5[0.3-0.9] |  | 0.3[0.1-0.9] |  | 15% (3) | 0.4[0.1-1.6] |  |  |  |
| **Currently attending school** |  |  |  |  |  |  |  |  |  |  |  |
| No | 18% (63) | 16% (21) | Reference | 0.482 |  |  | 5% (1) | Reference | 0.163 |  |  |
| Yes | 82% (281) | 84% (107) | 1.2[0.3–1.2] |  |  |  | 95% (18) | 4.2[0.6-32] |  |  |  |
| **History of Hospitalization for chest problems** |  |  |  |  |  |  |  |  |  |  |  |
| No | 98% (339) | 97% (124) | Reference | 0.154 | Reference | 0.195 | 95% (19) | Reference | 0.278 | Reference | - |
| Yes | 2% (6) | 3% (4) | 3.5[0.4 – 19] |  | 3.9[0.3-50] |  | 5% (1) | 3.4[0.4 – 30] |  | Omitted |  |
| **Ever treated for TB** |  |  |  |  |  |  |  |  |  |  |  |
| No | 72% (249) | 64%(81) | Reference | **0.006** | Reference | **0.015** | 80% (16) | Reference | 0.431 | Reference | 0.385 |
| Yes | 28% (95) | 36% (46) | 2.0 [1.2 -3.2] |  | 2.0[1.1-3.6] |  | 20% (4) | 0.6[0.2-2.0] |  | 0.6[0.2-2.0] |  |
| **Ever treated for asthma** |  |  |  |  |  |  |  |  |  |  |  |
| No | 97% (277) | 96% (111) | Reference | 0.359 | Reference | 0.778 | 6% (1) | Reference | 0.514 | Reference | 0.267 |
| Yes | 3% (9) | 4% (5) | 1.9[0.5-7.1] |  | 1.2[0.3-5.3] |  | 94% (16) | 2.0[0.2-17] |  | 3.8[0.2-2.0] |  |
| **Duration on ART (years)** |  |  |  |  |  |  |  |  |  |  |  |
| > 6 months to <2 years | 8% (28) | 12% (15) | Reference | 0.064 | Reference | 0.100 | 5% (1) | Reference | 0.693 | Reference | 0.702 |
| 2 to <4 years | 18% (60) | 23% (29) | 0.7[0.3- 1.8] |  | 1.4[0.5-4.3] |  | 15% (3) | 1.4[0.1- 14] |  | 1.7[0.1-35] |  |
| 4 to < 6years | 20% (67) | 20% (25) | 0.6[0.3 - 1.5] |  | 0.5[0.2-1.4] |  | 30% (6) | 2.7[0.3- 23] |  | 5.7[0.3-103] |  |
| 6years or more | 54% (183) | 45% (56) | 0.4[0.2 - 0.8] |  | 0.5[0.2-1.4] |  | 50% (10) | 1.6[0.3- 12] |  | 3.2[0.2-53] |  |
| CD4 count (Cells/ml) |  |  |  |  |  |  |  |  |  |  |  |
| < 200 | 11% (39) | 12% (16) | Reference | 0.591 |  |  | 15% (3) | Reference | 0.850 |  |  |
| 200-500 | 28% (98) | 29% (37) | 0.9[0.4–1.9] |  |  |  | 25% (5) | 0.6[0.1-2.8] |  |  |  |
| >500 | 60% (208) | 59% (75) | 0.8[0.4-1.6] |  |  |  | 60% (12) | 0.7[0.2- 2.7] |  |  |  |
| Baseline viral load |  |  |  |  |  |  |  |  |  |  |  |
| Unsuppressed | 45% (154) | 45% (57) | Reference | 0.871 | Reference | 0.866 | 65% (13) | Reference | 0.096 | Reference | 0.075 |
| Suppressed | 55% (191) | 55% (70) | 0.9[0.6-1.5] |  | 1.0[0.6-1.8] |  | 35% (7) | 0.4[0.2- 1.2] |  | 0.3[0.1-1.0] |  |
| ART regimen |  |  |  |  |  |  |  |  |  |  |  |
| NNRT-base-1st line | 77% (266) | 74% (95) | Reference | 0.328 | Reference | 0.596 | 75% (15) | Reference | 0.818 | Reference | 0.805 |
| PI-base 2nd line | 23% (79) | 26% (33) | 0.8 [0.5-1.3] |  | 1.2[0.6-2.2] |  | 25% (5) | 1.1[0.4 -3.2] |  | 1.2[0.3- 4.2] |  |
| Cotrimoxazole prophylaxis |  |  |  |  |  |  |  |  |  |  |  |
| No | 10% (33) | 10% (13) | Reference | 0.77 | Reference | 0.083 | 10% | Reference | 0.963 | Reference | 0.673 |
| Yes | 90% (307) | 90% (113) | 0.9[0.4–1.9] |  | 0.5[0.2-1.1] |  | 90% (18) | 1.0[0.2-4.4] |  | 1.6[0.2-14] |  |
| Baseline weight-for-age-z score |  |  |  |  |  |  |  |  |  |  |  |
| Not underweight | 55% (189) | 49% (63) | Reference | 0.552 |  |  | 50% (10) | Reference | 0.658 |  |  |
| Underweight | 45% (156) | 51% (65) | 1.4[0.9–2.2] |  |  |  | 50% (10) | 1.2[0.5-3.0] |  |  |  |
| Baseline height-for-age-z score |  |  |  |  |  |  |  |  |  |  |  |
| Not stunted | 54% (188) | 49% (63) | Reference | 0.131 | Reference | 0.141 | 60% (12) | Reference | 0.611 | Reference | 0.673 |
| Stunted | 46% (157) | 51% (65) | 1.4[0.9–2.2] |  | 0.5[0.2-1.1] |  | 40% (8) | 0.8[0.3-2.0] |  | 0.8[0.3-2.3] |  |

Abbreviations: NNRT = Nonnucleoside reverse transcriptase inhibitor, No. = number of participants, IQR = interquartile range, TB = tuberculosis, PI = Protease inhibitor, FEV1: Forced expiratory volume in one second;
Participants with missing responses were excluded from that variable: *currently attending school*, n=1; *antiretroviral regimen*, n=1; *history of TB*, n=1; *current cough*, n=1; *cotrimoxazole prophylaxis*, n=5; *viral load*, n=2; and *duration on ART*, n=7.
a Variables included in the multivariate logistic regression are participant group, age category, duration of ART, site, sex, height-for-age-z score, viral suppression, history of TB treatment, MRC dyspnea score and ART regimen

**Table 5.** Univariate and multivariate analysis of factors associated with nasopharyngeal HI and MC colonization at baseline.

| Variable | Total observations<br>(N = 345) | HI colonization |  |  |  |  | MC colonization (n= 84) |  |  |  |  |
| --- | --- | --- | --- | --- | --- | --- | --- | --- | --- | --- | --- |
|  |  | Colonized %<br>(n =143) | OR [95% CI] | *p* | Adjusted OR<br>[95% CI] | *p* | Colonized %<br>(n=84) | OR [95% CI] | *P* | Adjusted OR<br>[95% CI] | *P* |
| **Group** |  |  |  |  |  |  |  |  |  |  |  |
| HCLD- | 17% (58) | 13% (19) | Reference |  | Reference |  | 11% (9) | Reference |  | Reference |  |
| HCLD+ | 83% (287) | 87% (124) | 1.6 [0.9-2.8] | 0.143 | 2.4 [0.5- 10.3] | 0.248 | 89% (75) | 1.9[0.9 - 4.1] | 0.074 | 0.9[0.1-9.9] | 0.929 |
| **Sex** |  |  |  |  |  |  |  |  |  |  |  |
| Male | 48% (165) | 47% (67) | Reference |  | Reference |  | 45% (38) | Reference |  | Reference |  |
| Female | 52% (180) | 53% (76) | 1.1 [0.5-1.6] | 0.761 | 1.2 [0.7-1.9] | 0.579 | 55% (46) | 1.1[0.7 - 1.9] | 0.585 | 1.5[0.8-2.9] | 0.206 |
| **Study site** |  |  |  |  |  |  |  |  |  |  |  |
| Malawi | 27% (93) | 35% (50) | Reference |  | Reference |  | 33% (28) | Reference |  | Reference |  |
| Zimbabwe | 73% (252) | 65% (93) | 0.5[0.3-0.8] | **0.005** | 0.5 [0.3 – 0.9] | **0.014** | 67% (56) | 0.7[0.4 - 1.1] | 0.136 | 1.3[0.6-3.0] | 0.542 |
| **Baseline age category (years)** |  |  |  |  |  |  |  |  |  |  |  |
| 6 – 12 | 26% (91) | 29% (41) | Reference |  | Reference |  | 13% (11) | Reference |  | Reference |  |
| 13 – 16 | 41% (141) | 43% (62) | 1.0[0.6- 1.6] | 0.275 | 0.9[0.5-1.9] | 0.425 | 45% (38) | 0.7[0.3-1.3] | 0.326 | 1.2[0.6-2.4] | 0.839 |
| 17 – 19 | 33% (113) | 28% (40) | 0.7[0.4-1.2] |  | 0.6[0.3- 1.3] |  | 26% (22) | 1.0[0.6-1.9] |  | 1.4[0.6-3.4] |  |
| **Currently attending school** |  |  |  |  |  |  |  |  |  |  |  |
| No | 18% (63) | 16% (23) | Reference |  |  |  | 10% (8) | Reference |  | Reference |  |
| Yes | 82% (281) | 84% (120) | 1.3[0.7-2.3] | 0.368 |  |  | 90% (76) | 2.5[1.2-5.6] | **0.011** | 2.5[1.0-6.2] | **0.047** |
| **History of Hospitalization for chest problems** |  |  |  |  |  |  |  |  |  |  |  |
| No | 98% (339) | 98% (140) | Reference |  | Reference |  | 96% (81) | Reference |  | Reference |  |
| Yes | 2% (6) | 2% (3) | 1.4[0.3-7.1] | 0.670 | 1.9 [0.3 -13.8] | 0.529 | 4% (3) | 3.2[0.616] | 0.171 | 3.6[0.5-23] | 0.186 |
| **Ever treated for TB** |  |  |  |  |  |  |  |  |  |  |  |
| No | 72% (249) | 66% (94) | Reference |  | Reference |  | 67% (56) | Reference |  | Reference |  |
| Yes | 28% (95) | 34% (48) | 1.7[1.1-2.7] | **0.029** | 2.1 [1.2-3.9] | **0.009** | 33% (28) | 1.4[0.8-2.4] | 0.185 | 1.4[0.7-2.7] | 0.328 |
| **Ever treated for Asthma** |  |  |  |  |  |  |  |  |  |  |  |
| No | 97% (277) | 95% (118) | Reference |  | Reference |  | 96% (72) | Reference |  | Reference |  |
| Yes | 3% (9) | 5% (6) | 2.7[0.7-11] | 0.167 | 2.1 [0.5-9.6] | 0.322 | 4% (3) | 1.4[0.3-5.8] | 0.624 | 1.1[0.2-5.2] | 0.903 |
| **Duration on ART (years)** |  |  |  |  |  |  |  |  |  |  |  |
| > 6 months to <2 years | 8% (28) | 14% (20) | Reference | **0.005** | Reference | **0.007** | 14% (12) | Reference | **0.021** | Reference | **0.024** |
| 2 to <4 years | 18% (60) | 19% (26) | 0.3[0. -0.8] |  | 0.3[0.1-1.1] |  | 16% (13) | 0.4[0.1-1.0] |  | 0.5[0.1-1.6] |  |
| 4 to < 6years | 20% (67) | 20% (28) | 0.3[0.1-0.7] |  | 0.2[0.1-0.8] |  | 16% (13) | 0.3[0.1-0.8] |  | 0.3[0.1-1.0] |  |
| 6years or more | 54% (183) | 47% (65) | 0.2[0.1-0.5] |  | 0.3[0.1-0.9] |  | 54% (44) | 0.4[0.2-1.0] |  | 0.6[0.2-1.7] |  |
| **CD4 count (Cells/ml)** |  |  |  |  |  |  |  |  |  |  |  |
| < 200 | 11% (39) | 16% (23) | Reference | 0.065 |  |  | 15% (13) | Reference | 0.293 |  |  |
| 200-500 | 28% (98) | 27% (38) | 0.4[0.2-0.9] |  |  |  | 24% (20) | 0.5[0.2-1.2] |  |  |  |
| >500 | 60% (208) | 57% (82) | 0.5[0.8-2.7] |  |  |  | 61% (51) | 0.6[0.3-1.4] |  |  |  |
| **Baseline viral load** |  |  |  |  |  |  |  |  |  |  |  |
| Unsuppressed | 45% (152) | 45% (64) | Reference |  | Reference |  | 43% (35) | Reference |  | Reference |  |
| suppressed | 55% (191) | 55% (78) | 0.9 [0.6-1.5] | 0.813 | 1.0 [0.6-1.7] | 0.992 | 57% (48) | 3.3[0.2-54.8] | 0.664 | 1.3[0.7-2.3] | 0.436 |
| **ART regimen** |  |  |  |  |  |  |  |  |  |  |  |
| NNRT-base-1st line | 77% (266) | 78% (111) | Reference |  | Reference |  | 74% (62) | Reference |  | Reference |  |
| PI-base 2nd line | 23% (79) | 22% (32) | 1.0[0.6-1.6] | 0.846 | 0.9[0.5-1.7] | 0.737 | 26% (22) | 1.3[0.7-2.2] | 0.414 | 1.5[0.7-2.3] | 0.436 |
| **Cotrimoxazole prophylaxis** |  |  |  |  |  |  |  |  |  |  |  |
| No | 10% (33) | 10% (14) | Reference |  | Reference |  | 10% (8) | Reference |  | Reference |  |
| Yes | 90% (307) | 90% (126) | 0.9[0.5 -2.0] | 0.878 | 0.7 [0.3 -1.8] | 0.652 | 90% (75) | 1.1[0.4-2.3] | 0.981 | 0.6[0.2-1.6] | 0.353 |
| **Baseline weight-for-age-z score** |  |  |  |  |  |  |  |  |  |  |  |
| Not underweight | 55% (189) | 56% (80) | Reference |  |  |  | 51% (43) | Reference |  |  |  |
| Underweight | 45% (156) | 44% (63) | 0.9[0.6-1.4] | 0.715 |  |  | 49% (41) | 1.2[0.7-2.0] | 0.446 |  |  |
| **Baseline height-for-age-z score** |  |  |  |  |  |  |  |  |  |  |  |
| Not stunted | 54% (188) | 51% (73) | Reference |  | Reference |  | 56% (47) | Reference |  | Reference |  |
| Stunted | 46% (157) | 49% (70) | 1.3[0.8-1.9] | 0.280 | 1.1 [0.6 - 1.8] | 0.777 | 44% (37) | 0.9[0.6-1.5] | 0.757 | 0.8[0.5-1.5] | 0.524 |

Abbreviations: NNRT = Nonnucleoside reverse transcriptase inhibitor, No. = number of participants, IQR = interquartile range, TB = tuberculosis, PI = Protease inhibitor, FEV1: Forced expiratory volume in one second;
Participants with missing responses were excluded from that variable: *currently attending school*, n=1; *antiretroviral regimen*, n=1; *history of TB*, n=1; *current cough*, n=1; *cotrimoxazole prophylaxis*, n=5; *viral load*, n=2; and *duration on ART*, n=7.
a Variables included in the multivariate logistic regression are participant group, age category, duration of ART, site, sex, height-for-age-z score, viral suppression, history of TB treatment, MRC dyspnea score and ART regimen

## DISCUSSION

In this study, we used quantitative PCR to determine the prevalence and density of bacterial and viral carriage in HIV-infected African children. As previously shown [14], microbial colonization was more frequently detected in HCLD+ than HCLD-participants, with the former more likely to carry SP or HRV. Strikingly, viruses (predominantly HRV) were detected only in HCLD+ children. Moreover, we observed that HCLD+ participants had a higher HI and MC density than their HCLD-counterparts. The prevalence and densities of all SP serotypes tested were similar between the two groups, with more of the recovered SP serotypes (79%) being non-PCV 13. Study participants with a history of previous tuberclosis treatment were more likely to carry SP or HI, while those who used ART for ≥2 years were less likely to carry HI and MC. Furthermore, female study participants were less likely to carry SA, and those living in Zimbabwe were less likely to carry HI.

The prevalence of HI in the current study in both HCLD+ (43%) and HCLD-(33%) participants was higher than that observed in our previous study of the same cohort by Abotsi *et al*. [14] (12% and 5%, respectively). Similar studies conducted in India [29] and Zambia [30] in HIV-infected children also observed a lower prevalence (26% and 29%, respectively) than our current study (33%). The discrepancy in results could be attributed to the more sensitive PCR detection method used in our study compared to the culture method employed in previous studies.

Furthermore, HCLD+ participants showed a higher density of HI than their counterparts. Previous studies have associated HI carriage in participants with other lung diseases, including asthma [31], bronchiectasis [32, 33] and chronic obstructive pulmonary disease [34–36]. HI has been identified as a biomarker for predicting the response to azithromycin treatment in adults with persistent uncontrolled asthma [31]. It has also been associated with negative outcomes in children suffering from respiratory viral infections [37], including hospitalization among RSV-positive children [38]. The bacterium’s ability to invade host epithelial cells, evade host defense mechanisms, form biofilms and survive as an intracellular pathogen contributes to its pathogenic nature [39], which may suggest an important role that it may play in HCLD+ pathogenesis. However, further studies are needed to further elucidate this observation.

The prevalence of carriage of SA in this study (HCLD+ [6%] and HCLD-[5%]) was markedly lower than that observed using bacterial culture in the same cohort (HCLD+ [23%] and HCLD-[19%]) [14]. This difference in prevalence may be related to the efficiency of the nucleic acid extraction method used. The extraction of nucleic acids requires extended and vigorous lysis steps for some bacterial species (gram-positive such as SA) compared to others (gram-negative such as HI and MC) [40]; however, for this study, although we used a rigorous extraction protocol incorporating bead-beating, the inherent complexity and resilience of the SA bacterial cell wall may have contributed to the low yield. This underscores the need for tailored approaches and ongoing refinement of extraction methods to improve yield consistency. Additionally, carriage may have been influenced by the efficiency of the annealing of PCR primers [41]. Therefore, further research on our cohort should involve blasting the primers used in this study against the SA genome to confirm their effectiveness in targeting all the SA strains accurately.

High SP, HI and MC density in the nasopharynx has been associated with respiratory infections in children [42, 43]. This is consistent with our study, where a higher HI and MC density was observed in HCLD+ participants than in their HCLD-counterparts. The HCLD+ participants may have chronic lung infection, as evidenced by the isolation of bacteria from their sputum in our previous study [14]. Microbiota dominated by *Haemophilus*, *Moraxella* or *Neisseria* species are associated with chronic lung diseases, including chronic obstructive pulmonary disease and asthma [44–47]. Bhadriraju *et al.* [39] observed that HIV-infected children with a sputum bacteriome dominated by *Haemophilus*, *Moraxella* or *Neisseria* species were 1.5 times more likely to have HCLD than those with *Streptococcus* or *Prevotella* spp. [39]. These bacterial genera were also associated with enhanced inflammatory effects [39]. Interestingly, we detected *Neisseria* species (*N. lactamica*) in HCLD+ participants only. Taken together, these findings support the important role of HI and MC in HCLD. Our observation of a higher SP carriage in the HCLD+ group than in the HCLD- group is consistent with our culture-based study of the same cohort [14]. Furthermore, SP carriage in the HCLD-participants (21%) is comparable to studies of HIV-infected children in South Africa (22.2%) [48] and Cambodia (17.6%) [41, 49]. Nevertheless, the prevalence is higher than that observed in children living with HIV in Ethiopia (10.3%) [50] and lower than that in studies from Ghana (27.1%) and Tanzania (81%) [51]. The differences in SP prevalence observed between studies can again be related to differences in the age of participants—younger children have a higher carriage prevalence[30, 51], socioeconomic factors [52] and the geographical location of the participants.

The prevalence of PCV 13 serotypes and densities in both study groups (HCLD+: 30.4% and HCLD-: 16.7%) did not differ significantly. The most prevalent PCV-13 serotypes were serotypes 4 (15.9%), 19F (15.9%), 19A (15.9%) and 18C (14%). A study of HIV-infected children in Malawi [53] reports 19F and 6A among the most predominant serotypes. The relatively high prevalence of serotype 19A in PCV-vaccinated children has been suggested by Kamng’ona *et al.* [53] to occur due to an inversion in the *rmlD* gene at the CPS locus. This may downregulate the *rmlD* gene on the CPS locus, causing an altered 19A capsule [54] that is not recognized by the PCV vaccine.

There was a higher prevalence of non-PCV13 serotypes in both the HCLD+ (70.7% [99/140]) and HCLD-(83.3% [15/180]) compared to PCV13 serotypes. Similar findings were reported in Malawi [53], Nigeria [55] and Ghana [56].. We assume that community herd protection from vaccinated siblings, neighbors, and playmates may be responsible for the low prevalence of vaccine-type serotypes in our cohort. Continued surveillance of SP and its non-PCV 13 serotypes is warranted to inform future vaccine formulation and roll-out strategies, especially in this vulnerable population.

The major risk factors associated with the development of pneumococcal disease are demographic (age and sex) and immune status (CD4 count and HIV viral load) [57]. We observed no association between these common factors and most bacterial species, including SP. This is supported by previous studies that reported a lack of association between CD4 count and the prevalence of pneumococcal carriage [30, 58, 59]. A longer period on an ART regimen (two years or more) was associated with reduced carriage of MC and HI. Similar results were obtained from a study among HIV-infected adults in Brazil [60]. ART therapy could help reduce the risk of infection and carriage through immune reconstitution [60].

The presence of viruses increases bacterial adherence, and the difference in the prevalence of viruses in HCLD+ *vs* HCLD-children may partially explain the increased HI and MC densities we observed. Our findings are consistent with a study by Binks *et al.* [43], who reported an increased SP and HI density during coinfection with respiratory viruses within the nasopharynx of Australian children with otitis media. However, no significant difference in the bacterial load was detected in SP from the HCLD+ and HCLD- groups. Viruses expose the host to bacterial infection through various mechanisms, including the destruction of the respiratory epithelium, modulation of innate defenses and alteration of cell membranes, which facilitates bacterial adherence [15]. Ishizuka *et al.* [61], in their *in vitro* studies, observed increased SP adherence to epithelial cells after infection with HRV. They suggested that this observation may explain why pneumonia develops following an HRV infection [61]. Interestingly, we found no association between any virus (HRV, RSVA and RSVB) and the prevalence or density of carriage of SP or other bacterial species tested. This contrasts with several *in vitro* and *in vivo* studies that have suggested that respiratory virus infection increases bacterial adherence and subsequent bacterial superinfection within the nasopharynx [61–63]. This discrepancy may be explained by the few viruses we detected due to the limited sample size, especially in the HCLD- group.

HRV is responsible for most upper respiratory tract infections and their complications, including bronchitis [15]. In a study of HIV-infected children in India [64], HRV was the most prevalent virus in these participants when asymptomatic. The GABRIEL multicenter case‒control study in Africa and Asia also found HRV in healthy control groups of pneumonia childhood studies [65]. In contrast, our study detected HRV (7%) in only HCLD+ participants. Notably, RSV infection was uncommon, consistent with previous studies conducted in Africa and Asia (PERCH case‒control studies [66] and DCHS case‒control studies [67]), which showed its infrequency except during acute respiratory infections.

In conclusion, our study findings indicate that HCLD+ participants were more commonly colonized by any of the bacteria tested compared to HCLD-participants. Specifically, the HCLD+ group had a higher prevalence of carriage of SP bacteria, as well as a higher density of MC and HI bacteria. Interestingly, viruses, particularly HRV, were detected only in the HCLD+ group. Moreover, our research revealed that previous treatment for tuberculosis was positively associated with carriage of HI or SP bacteria among study participants. On the other hand, being a female participant was found to be less likely to be associated with SA carriage. Additionally, longer periods on the ART regimen were associated with reduced carriage of HI or MC bacteria. Our study sheds light on the quantitative information on microbial carriage and nasopharyngeal carriage of viruses and serotypes of HI and SP in children with HCLD+. A limitation of our study is the small sample size of HCLD-participants, which may have affected the statistical power and generalizability of our findings. Therefore, there is need for more comprehensive studies in this population to further investigate the role of SP, HI, MC, and HRV in the pathogenesis of CLD and the underlying mechanisms behind these bacterial associations.

## Abbreviations

ART: antiretroviral therapy
AZM: azithromycin
HCLD: HIV-associated chronic lung disease
HCLD+: HIV-infected patients with HIV-associated chronic lung disease.
HCLD-: HIV-infected patients without HIV-associated chronic lung disease
NP: nasopharyngeal
HI: *Haemophilus influenzae*
MC: *Moraxella catarrhalis*
SA: *Staphylococcus aureus*
SP: *Streptococcus pneumoniae*
HRV: human rhinovirus
RSV: respiratory syncytial virus

## Acknowledgments

We would like to acknowledge the BREATHE trial participants, their families and the study team. We would also like to thank the staff of the Division of Medical Microbiology and the Department of Molecular and Cell Biology, particularly members of Dube Lab and the UCT community, for providing all the resources required for the project and training whenever needed. We are grateful to Dr. Courtney Olwagen and Lara Van Der Merwe of the WITS-VIDA research team for assistance with the Fluidigm assay and data analysis.

## Authors’ contributions

FSD conceived the study. PKM conducted the laboratory experiments and data analysis and wrote the first draft of the manuscript supervised by REA and FSD. JOO, MN and RAF conceived and led the parent BREATHE study. JOO secured funding from GLOBVAC on behalf of the consortium. All authors contributed to, read and approved the final manuscript.

## Funding

This parent study was funded by the Global Health and Vaccination Research (GLOBVAC) Programme of the Medical Research Council of Norway. This substudy was funded by the Royal Society through the Future Leaders African Independent Research award and the National Institute for Health Research (NIHR) Global Health Research Unit on Mucosal Pathogens using UK aid from the UK Government (Project number 16/136/46). The views expressed are those of the authors and not necessarily those of the NIHR, the Department of Health and Social Care. PM received funding from the UCT Postgraduate Funding, UCT’s Building Research Active Academic Staff (B.R.A.A.S.) award, the Molecular and Cell Biology_Equity Development Programme scholarship, and the Dube-lab scholarship. REA acknowledges the financial support of the Swedish International Development Cooperation Agency (SIDA) through the Organization of Women in Science for the developing world (OWSD) PhD Fellowship, Margaret McNamara Education Grants and L’Oréal UNESCO For Women in Science PhD Fellowship. MN is supported by an Australian National Health and Medical Research Council Investigator Grant [APP1174455]. FSD is supported by the National Research Foundation of South Africa (112160), Future Leaders – African Independent Research (FLAIR) Fellowship, the National Institute for Health Research (NIHR) Global Health Research Unit on Mucosal Pathogens using UK aid from the UK Government, the University of Cape Town and the Allergy Society of South Africa (ALLSA). RAF is funded by the Wellcome Trust (206316_Z_17_Z). The funders had no role in the design of the study and collection, analysis, and interpretation of data and in writing the manuscript.

## Availability of data and materials

The datasets used and analyzed during the current study are available from Felix Dube on reasonable request and ethical approval.

## Ethics approval and consent to participate

The parent study (BREATHE) was approved by the Human Research and Ethics Committee of the University of Cape Town - UCT HREC (HREC/REF:754/2015), the London School of Hygiene and Tropical Medicine Ethics Committee (reference 8818), the Harare Central Hospital Ethics Committee and Medical Research Council of Zimbabwe (reference MRCZ/A/1946), the College of Medicine Research Ethics Committee Malawi (reference P.04/15/1719) and the Regional Committee for Medical and Health Research Ethics, Northern Norway (reference 2015/1650). The University of Oxford waived approval. Additional ethical approval was received for this substudy from the UCT HREC (HREC/REF: 092/2019). No additional data were collected other than those approved in the parent study. Written informed consent and assent were given by guardians and participants, respectively. Participants who were 18 years old and above consented independently at the time of enrollment. All data obtained and generated during the study were kept confidential. This research was conducted in accordance with the Declaration of Helsinki.

## Consent for publication

Not applicable.

## Competing interests

The authors declare that they have no competing interests.

## Notes

### Competing Interest Statement

The authors have declared no competing interest.

### Clinical Trial

NCT02426112

### Author Declarations

The parent study (BREATHE) was approved by the Human Research and Ethics Committee of the University of Cape Town(HREC/REF:754/2015), the London School of Hygiene and Tropical Medicine Ethics Committee (reference 8818), the Harare Central Hospital Ethics Committee and Medical Research Council of Zimbabwe (reference MRCZ/A/1946), the College of Medicine Research Ethics Committee Malawi (reference P.04/15/1719) and the Regional Committee for Medical and Health Research Ethics, Northern Norway (reference 2015/1650). The University of Oxford waived approval. Additional ethical approval was received for this substudy from the UCT HREC (HREC/REF: 092/2019). No additional data were collected other than those approved in the parent study.

